# Occurrence, Risk factors and Antimicrobial resistance of *Campylobacter* from Poultry and Humans in Central Ethiopia: A One Health Approach

**DOI:** 10.1101/2025.02.14.25322269

**Authors:** Fikre Zeru, Haileeyesus Adamu, Yohannes Hagos Woldearegay, Tesfaye Sisay Tessema, Ingrid Hansson, Sofia Boqvist

## Abstract

**Background:** *Campylobacter* is a leading foodborne pathogen posing a significant One Health challenge due to its broad animal reservoirs and serious antibiotic resistance issues. Despite frequent human-animal-environment interactions in Ethiopia, One Health studies on the occurrence and transmission of *Campylobacter* are crucial but lacking.

**Methodology/Principal Findings:** A cross-sectional study from March 2021 to March 2022 in and around Debre Berhan, Ethiopia, examined *Campylobacter* occurrence, resistance, and risk factors in poultry and humans using a One Health approach. A total of 366 samples from 122 poultry farms were collected, including cloacal swabs, human stools, and poultry house floor sock samples. Epidemiological data on risk factors and respondents awareness were gathered through face-to-face interviews. *Campylobacter* spp. were isolated following ISO 10272 and confirmed with multiplex PCR, with antimicrobial susceptibility tested by disc diffusion according EUCAST guidelines. *Campylobacter spp.* were found in 12.5% of samples, highest in poultry (19.6%), followed by human stools (13.1%) and floor socks (4.9%). *Campylobacter jejuni* was the dominant species (80.4%), followed by *C. coli* (19.6%). In poultry, mixed farming with cattle increased *Campylobacter* colonization odds (adjusted odds ratio; AOR=9.5), while all-in/all-out management decreased it (AOR=8.4). In humans, *Campylobacter* infection was linked to raw milk consumption (AOR=5.5), poultry access to living areas (AOR=6.3), not using personal protective equipment working with poultry (AOR=8.3) and not washing hands after handling poultry and cleaning barn (AOR=5.6). Farm workers had a significant knowledge gap on zoonotic risks, including *Campylobacter* and One Health. High antibiotic resistance was observed, especially to erythromycin (63.0%), ciprofloxacin (69.5%), tetracycline (89.1%), and oxytetracycline (73.9%), with 69.5% of isolates showing multi-drug resistance.

**Conclusions/Significance:** The study revealed widespread occurrence of resistant *Campylobacter* spp. in poultry, workers, and the environment, highlighting the need for One Health interventions, including better biosecurity, hygiene, education, and stricter antimicrobial use to safeguard animal and human health.

**Author summary:** A study conducted in and around Debre Berhan, central Ethiopia, analyzed samples from poultry farms for *Campylobacter* spp. and their antimicrobial resistance. The highest prevalence of *Campylobacter* was found in poultry, followed by human and poultry house floor samples. Poor farm biosecurity and management practices were linked to *Campylobacter* in poultry, while human infections were associated with raw milk consumption and inadequate hygiene practices in poultry farm. The study emphasizes the zoonotic risks of *Campylobacter* and the need for a One Health approach to address its spread. *Campylobacter* isolates showed high resistance to common antimicrobials, with many classified as multidrug-resistant. Co-occurrence of *C. jejuni* in poultry, farm workers, and the environment, all with similar multidrug-resistant patterns, suggests possible transmission between them. These findings underline the widespread antimicrobial resistance in *Campylobacter* from poultry farms and the urgent need for responsible antibiotic use to control resistant strains. Given the serious implications of antimicrobial resistance, the zoonotic importance of *Campylobacter*, and the frequent human-animal-environment interactions in Ethiopia, it is crucial to implement a national plan for surveillance, prevention, and control, and along with promoting rational antimicrobial use through a One Health approach

## Introduction

*Campylobacter* is the top foodborne pathogen associated with animal-source food products globally and an important zoonotic pathogen [1]. The rising global incidence of *Campylobacter* infections underscores the urgent need for greater attention to this growing health burden [2]. *Campylobacter jejuni* and *Campylobacter coli* have extensive animal reservoirs, commonly colonizing the intestines of food-producing and companion animals [3]. Poultry serves as a major reservoir for *Campylobacter* spp. and is a significant source of transmission to humans [4]. Infections also occur through direct contact with animal feces, through the environment and food products other than poultry [4, 5]. The wide distribution, multiple transmission routes, and serious antibiotic-resistance [4] make *Campylobacter* a significant One Health concern.

*Campylobacter* is widespread in poultry farm environments, making farm management and biosecurity crucial for preventing transmission. Several risk factors for *Campylobacter* in broiler flocks have been identified, i.e., proximity to other livestock, poor general tidiness at the farm, insufficient hygiene and biosecurity practices, old broiler houses, and high broiler age at slaughter [6, 7].

Globally, the proportion of antimicrobials showing resistance above 50% increased from 0.15 to 0.41 in chickens [8]. Resistant *Campylobacter* strains have been recognized as a serious public health threat by global stakeholders such as the WHO and CDC [9]. Due to its zoonotic nature, *Campylobacter* is exposed to antibiotics used in both human healthcare and veterinary practices, presenting a clear and imminent challenge to the One Health approach. The bacterium rapidly evolves under antibiotic pressure, adapting to various hosts and environments, which leads to multidrug-resistant variants. Additionally, *Campylobacter* can acquire antimicrobial resistance (AMR) determinants from other bacteria through horizontal gene transfer [10]. Although *Campylobacter* typically does not cause clinical signs in animals, antibiotics are often used prophylactically in animal production. This irresponsible, unregulated antibiotic use is a major driver of AMR and contributes to the development of resistant strains, complicating the treatment of human infections [11]. Global data on *Campylobacter* shows that in low- and middle-income countries, the highest resistance rates are found for tetracycline (60%) and quinolones (60%) (8). Studies from various parts of Ethiopia report antimicrobial resistance rates for *Campylobacter* species ranging from 0–100% [12].

Poultry farming is a vital sector in Ethiopia’s agriculture, with approximately 57 million chickens, predominantly indigenous breeds in rural Ethiopia and commercial improved breeds farming in urban and peri-urban areas of central Ethiopia [13]. However, challenges like disease management, limited feed access, and low technical knowledge affect productivity [14]. Few studies have been published regarding *Campylobacter* prevalence in humans, animals, and food in Ethiopia [12]. Despite its critical role as a zoonotic pathogen, understanding the prevalence and transmission dynamics of *Campylobacter* in poultry farms in Ethiopia is limited. With frequent interactions among humans, animals, and the environment, comprehensive One Health studies are aslo essential but still insufficient. This study aimed to assess the occurrence of *Campylobacter* in poultry and humans in central Ethiopia, determine its antimicrobial resistance profile, and assess risk factors, farm worker awareness, and practices related to its spread using One Health approach.

## Materials and methods

### Study area and setting

The study was conducted in and around Debre Berhan town, central Ethiopia (Fig 1), approximately 120 km northeast of Addis Ababa. Debre Berhan serves as the capital, administrative, and economic center of the North Shewa Zone in the Amhara region. The area experiences a typical subtropical highland climate, characterized by two distinct seasons: the wet season, from June to September, and the dry season, from October to May. The annual humidity in the area is recorded at 62.7% [15]. The local farming system in the area is a mixed system, involving both crop cultivation and livestock rearing. Poultry production plays a significant role in the region, serving as an important source of meat and eggs for the local population. Like other parts of central Ethiopia [14], commercial poultry farming is growing in the urban and peri-urban areas of Debre Berhan.

**Fig 1:**
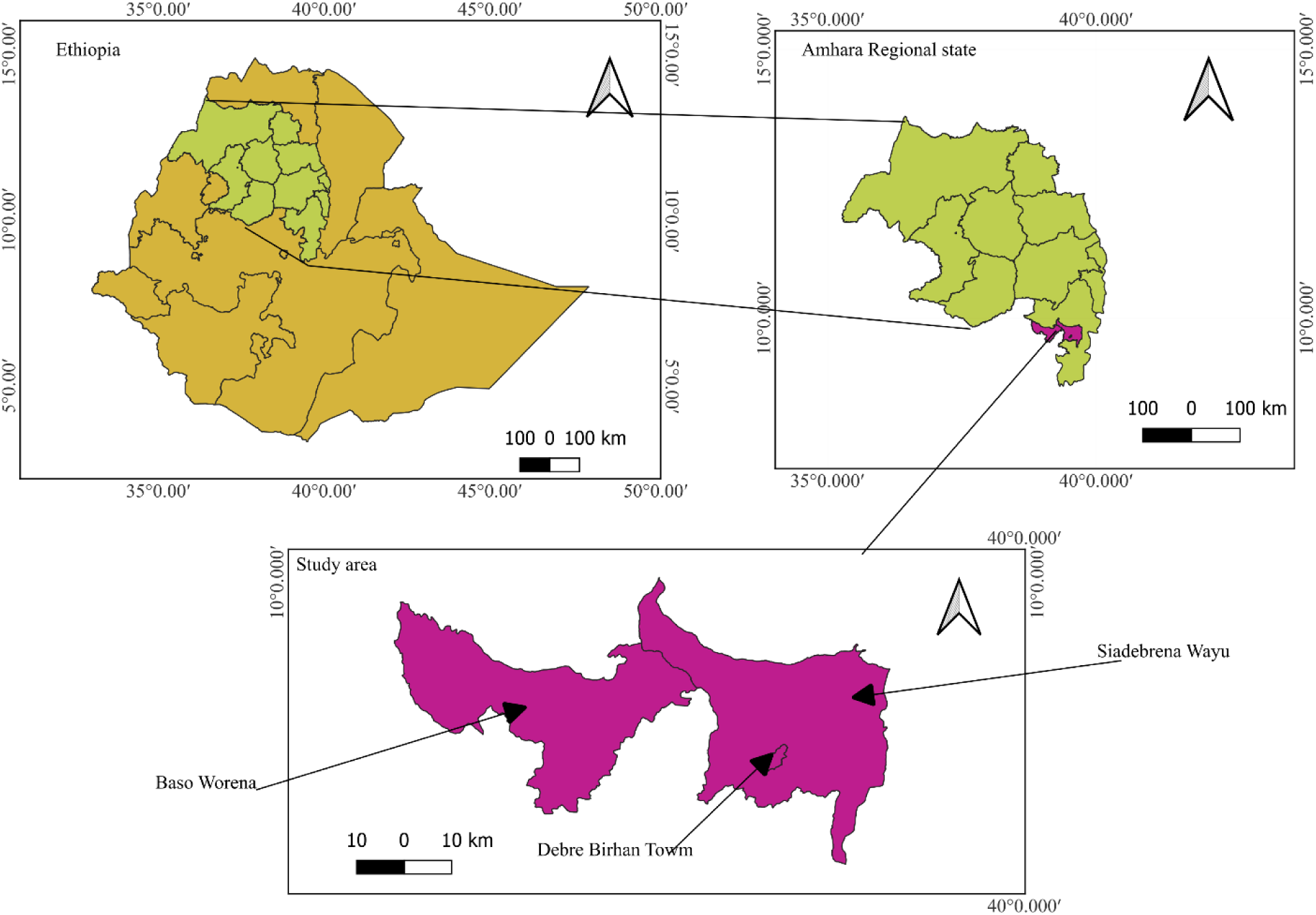
Map of the study area - Debre Berhan town, Siaderena Wayu and Baso Worena Woreda (generated using Quantum Geographic Information System (QGIS) version 3.34.12).

### Study approach

A cross-sectional study was conducted from March 2021 to March 2022 on selected poultry farms, employing a One Health approach that integrated humans, animals, and the environment. The study districts were chosen due to their high concentration of poultry farms, including Debre Berhan town, Basona Worena, and Siyadebrina Wayu. Before the study commenced, visits were made to the Agricultural Offices in each of the selected districts to obtain approval and gather information on the list of poultry farms, which was then used as the sampling frame. A veterinary professional in each district, who was familiar with the locations, was enlisted to assist in identifying poultry farms for inclusion in the study. The sample size was calculated using EpiInfo™ version 7.2.5.0 (Centers for Disease Control and Prevention, USA), with an expected prevalence of 50%, a 95% confidence level, a 5% absolute precision, and a design effect of 1.0. According to the Agricultural Office, there were 159 poultry farms in the study area, which were used to apply the finite population correction. Initially, 113 poultry farms were planned for selection; however, the sample size was increased to 122 to enhance the statistical power and reliability of the study. The main inclusion criteria for poultry farms were accessibility and willingness of the farmers to participate in the study. One sample was collected from each One Health domain at each farm. Specifically, three samples were obtained from each farm: one chicken sample (randomly chosen), one human sample (selected through a lottery method if more than one human was present), and environmental samples from all compartments of the poultry house floor (Fig. 2).

**Fig. 2:**
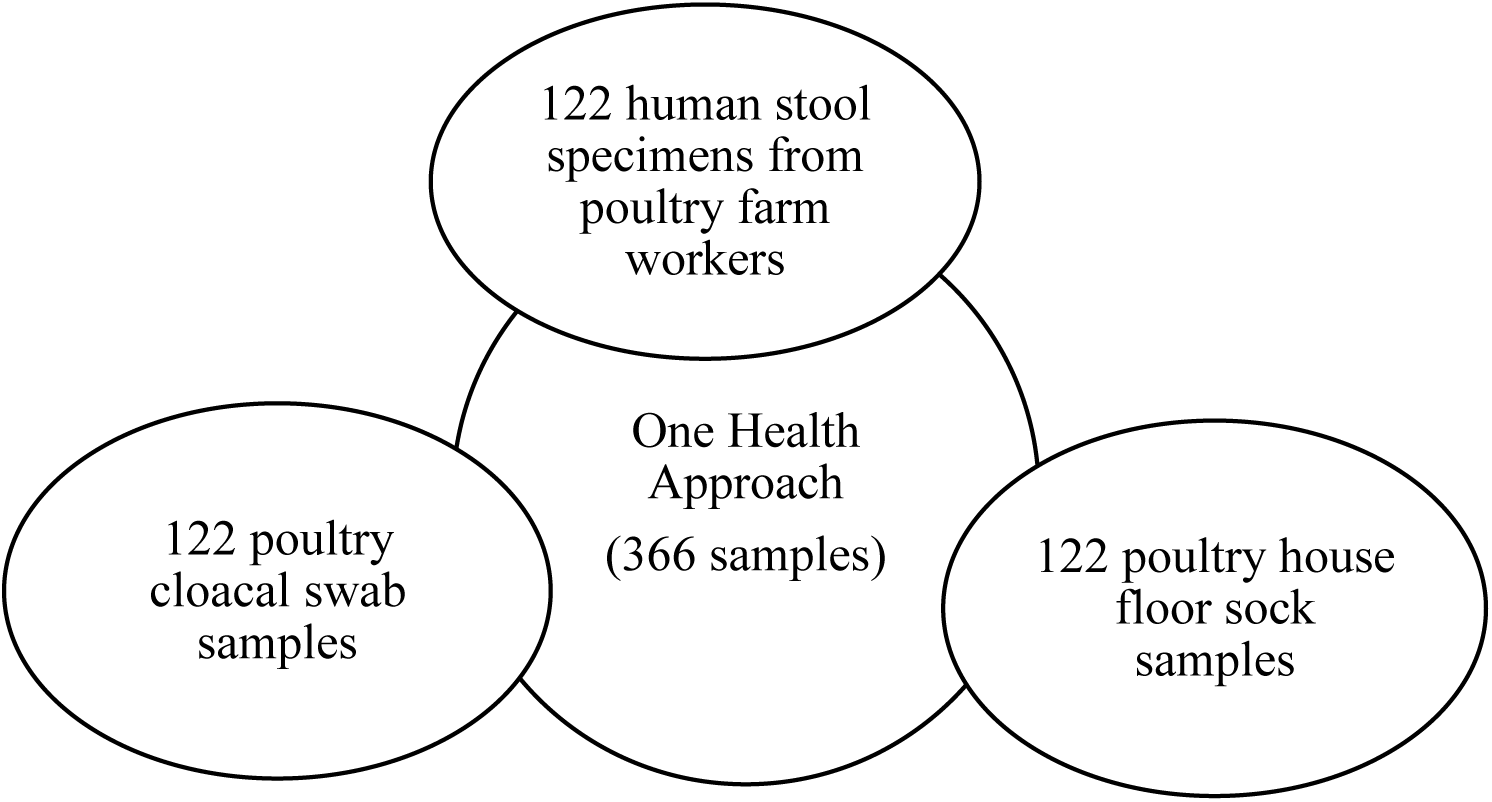
One Health approach to sample collection with distribution of the collection of 366 samples from poultry, humans, and the environment on selected farms in a study investigating occurrence of *Campylobacter* in and around Debre Berhan Ethiopia.

### Data collection

#### Epidemiological data collection

Epidemiological data was collected using a structured questionnaire designed to assess potential risk factors associated with *Campylobacter* infection and its transmission, and the respondent’s general awareness of zoonotic diseases. The questions included a mix of open, closed, and semi-closed formats. The questionnaire was organized into four sections that address farm practices associated with *Campylobacter* risk factors for poultry; consumption habits, occupation, and hygiene practices related to human *Campylobacter* infection; poultry house conditions, and awareness of poultry farm workers regarding *Campylobacter* as a zoonosis and the One Health concept. The questionnaire was administered through face-to-face interviews, translated orally into the local language (Amharic) during the interviews to facilitate better understanding. It was pretested on ten farms, and adjustments were made to improve clarity and relevance.

### Sample collection and transportation

Human stool specimens were collected from individuals working on the selected farms who had the most direct contact with poultry. Approximately 10 grams of self-collected human stool specimens were placed into sterile 50 ml screw-capped jars containing Cary-Blair Transport Medium (Oxoid, Basingstoke, UK). Individuals were given a few hours to provide the sample if unable to provide it immediately. Cloacal swab samples were collected from randomly selected poultry on the farm by a veterinarian using sterile cotton swabs, which were gently inserted into the poultry’s cloaca. Immediately after sampling, the cotton swabs were transferred into Amie’s agar gel with charcoal (Copan Diagnostics, Inc., Murrieta, CA, USA) for transport to the laboratory. Sock samples were collected from the poultry farm floor using boot sock sampling, as described by Hansson et al. [17]. Sterile boot socks, moistened with Cary-Blair transport medium, were worn over the footwear, and the sampler walked through all areas of the farm floor multiple times to ensure comprehensive sampling. After sampling, each sock was placed into a clean plastic bag with 10 ml of Cary-Blair Transport Medium, then securely sealed and labeled for identification.

All collected samples were placed in an icebox with ice packs until fieldwork was completed (up to 6 hours). They were then stored in a refrigerator at +4°C at the veterinary clinic of the respective district for a maximum of 24 hours. Subsequently, the samples were shipped in an icebox with ice packs to the Health Biotechnology Laboratory of the Institute of Biotechnology (IOB), Addis Ababa University (AAU), which took 2–3 hours. Upon arrival at the laboratory, the samples were processed immediately.

#### Isolation and Identification of *Campylobacter* species

The analyses of *Campylobacter* were carried out using a modified version of ISO 10272-1:2017 [18]. Human stool and cloacal swab samples, expected to have high concentrations of *Campylobacter*, were directly plated onto modified Charcoal Cefoperazone Deoxycholate Agar (mCCDA) (Oxoid, Basingstoke, UK) supplemented with CCDA Selective Supplement (SR0155E, Oxoid, Thermo Scientific Inc, Basingstoke, UK). For sock samples, which were expected to have lower concentrations of *Campylobacter* and higher levels of background microflora, enrichment was performed in 90 ml of buffer peptone water without selective antibiotic supplement (HiMedia Laboratories, Mumbai, India) [19]. The enrichment culture was incubated in a microaerobic atmosphere generated by CampyGen^®^ (Oxoid, Basingstoke, UK) at 42°C for 24 hours. After incubation, two loopfuls of the enrichment culture were plated onto mCCDA. The mCCDA plates were incubated at 42°C for 48 hours in a micro-aerobic atmosphere. If no growth was observed, the plates were re-incubated under microaerobic conditions for an additional 24 hours, extending the total incubation time to 72 hours.

### Confirmation and species Identification

Typical or suspected *Campylobacter* colonies were selected for further confirmation and species identification. The selected colonies were streaked onto a non-selective agar plate (Oxoid, Basingstoke, UK) containing 5% sheep blood and incubated at 37°C for 44 hours under microaerobic conditions, provided by CampyGen**^®^**. DNA extraction was performed using a heat and snap-chilling method according to a previous study by Barakat et al. [20]. Two to three colonies of fresh bacterial growth were collected from the blood agar plate, suspended in nuclease-free deionized water, and then heated at 95°C for 10 minutes in a heat block. The samples were then cooled immediately and centrifuged at 13,000g for 5 minutes at room temperature. The supernatant, containing the template DNA, was carefully transferred to nuclease-free Eppendorf tubes and stored at −20°C until further use.

The Multiplex PCR (mPCR) method was applied for the confirmation of *Campylobacter* isolates, both at the genus and species level, as described in previous studies [21–24]. mPCR amplification was carried out in 25 μl reaction volumes using a Himedia Thermal Cycler. The reaction mixture contained the following components: 1 μl of template DNA, 25 μM of each primer, 1.5 mM MgCl₂, 1250 μM of each dNTP, 5 μM of the green fluorescent nucleic acid stain, 1× reaction buffer, and 1 U of Taq DNA polymerase. The optimal mPCR conditions were as follows: initial denaturation at 96°C for 3 minutes, followed by 35 cycles of 96°C for 30 seconds, 57°C to 62°C for 90 seconds, and 72°C for 30 seconds. A final extension step was carried out at 72°C for 10 minutes [25]. The PCR products were analyzed by electrophoresis on a 1.5% agarose gel in TAE running buffer (10X). Ethidium bromide (0.5 μg/ml) was used for staining, and DNA ladder along with 6x loading dye was applied for visualization during gel electrophoresis.

### Antimicrobial susceptibility testing

The antimicrobial susceptibility of isolated *Campylobacter* was tested using the disc diffusion method, following European Committee on Antimicrobial Susceptibility Testing (EUCAST) guideline [26]. Three to four morphologically pure bacterial colonies were picked and suspended in sterile normal saline. The bacterial suspension was standardized by comparing it to a 0.5 McFarland standard. A loopful of the suspension was streaked onto Mueller-Hinton agar (HiMedia) supplemented with 5% sheep blood and evenly spread across the plate using a sterile cotton-tipped applicator. Antibiotic discs were placed on the plate using sterile forceps, spaced 20 mm apart. The plates were then incubated under microaerobic conditions at 42°C for 24-48 hours. Inhibition zones were measured at 24 and 48 hours and interpreted according to EUCAST guidelines. The antibiotics were selected based on their availability and common use in Ethiopia, and belonged to five antibiotic classes: quinolones (nalidixic acid (30 µg), ciprofloxacin (5 µg), and norfloxacin (10 µg)), macrolides (erythromycin (15 µg)), aminoglycosides (streptomycin (10 µg) and gentamicin (10 µg)), phenicols (chloramphenicol (30 µg)), and tetracyclines (tetracycline (30 µg) and oxytetracycline (30 µg)). Multi-Drug Resistance (MDR) was defined as non-susceptibility to at least three antibiotics classes. Established breakpoints according to EUCAST guidelines were only available for ciprofloxacin, erythromycin, and tetracycline. Due to the absence of validated cut-off values for *Campylobacter* resistance to other antibiotics at the time of this study, only an inhibition zone of 0 mm was considered resistant.

### Data management and analysis

Data from field surveys and laboratory results were recorded using Microsoft Excel 2010 (Microsoft Corporation, USA). Analysis was performed using STATA version 18.0 software (StataCorp LLC, USA). Descriptive statistics were used to summarize the data. To examine the relationship between risk factors and the occurrence of *Campylobacter* in chickens and humans, univariable logistic regression analysis was conducted, with results expressed as crude odds ratios (COR) and 95% confidence intervals (CI). Variables with P < 0.20 in the univariable analysis were further included in the multivariable logistic regression analysis. Results are reported as adjusted odds ratios (AOR) with 95% CI. A step-wise approach was used for model development, and the goodness-of-fit of the final model was assessed using the Hosmer-Lemeshow test with 10 groups. Multicollinearity and interaction effects were checked. The model’s predictive power was evaluated using the receiver operating characteristic (ROC) curve, and a P-value below 0.05 was considered as statistical significance.

### Ethical considerations and approval

A formal letter of support was subsequently obtained from AAU-IOB to communicate with the Agricultural Offices in each of the selected districts. The purpose of the study was explained to all participants, emphasizing that their participation was voluntary, and that all data would be handled anonymously although the results derived from this study would be published. Informed verbal consent was obtained from all participants before the commencement of data collection.

The study received ethical approval from the Institute of Biotechnology Institutional Review Board (Ethical Approval no. IOB/LB/2016/2024).

## Results

### Description of the poultry farms

Of the 122 poultry farms visited in Debre Berhan town and surrounding areas, 63.9% were located in peri-urban, 22.1% in urban, and 13.9% in rural areas. The farming system consisted of 47.5% smallholder farms (with 100 or fewer poultry, using local feed and basic management practices) and 52.5% commercial farms. Of the farms, 57.4% kept layers, 22.9% growers, 14.7% broilers, and 4.9% were mixed (layer and broiler). A significant proportion (72.9%) of farms lacked basic biosecurity measures, such as footbaths/changing shoes at the poultry house gate and disposed of dead poultry in garbage or open air near the poultry house. Additionally, 42.6% did not restrict access to unauthorized personnel, 45.9% did not separate sick poultry, and 64.7% used window ventilation. Furthermore, 66.4% of farms kept other animals, such as cattle, small ruminants, pets, and swine. The use of antibiotics, mainly oxytetracycline, as prophylactics was reported in 43.4% of the poultry farms visited.

#### Occurrence of *Campylobacter*

*Campylobacter* spp. were detected in 46 (12.5%) of 366 samples, with the highest prevalence in poultry (19.6%), followed by human samples (13.1%) and poultry house floor socks (4.9%). Of the 46 *Campylobacter* isolates, 37 (80.4%) were identified as *C. jejuni* and 9 (24.3%) as *C. coli* (Table 1). *Campylobacter* spp. were found in at least one sample from 39 (32.0%) of 122 poultry farms. On these farms, *Campylobacter* was found only in human stool on 12 (9.8%) farms, only in cloacal swabs on 18 (14.8%) farms, and only in floor sock samples on 3 (2.5%) farms. Moreover, *C. jejuni* was isolated from multiple sample types on 6 (4.9%) farms. Specifically, *C. jejuni* was detected in both human stool and cloacal swab in three farms, in both cloacal swab and floor sock samples in two farms, and in human stool, cloacal swab, and floor sock samples in one farm.

**Table 1.** Occurrence and distribution of *Campylobacter* spp. across sample types from poultry farms in and around Debre Berhan town, Ethiopia (n=366).

| Sample type | No. of samples | Positive |  |  | OR |  |
| --- | --- | --- | --- | --- | --- | --- |
|  |  | *C. jejuni* (%) | *C. coli* (%) | Total (%) | 95% CI | P-value |
| Cloacal swabs | 122 | 17 (13.9) | 7 (5.7) | 24 (19.6) | 4.7 (1.9-12.0) | 0.001 |
| Human stool | 122 | 14 (11.5) | 2 (1.6) | 16 (13.1) | 2.9 (1.1-7.7) | 0.031 |
| Sock samples | 122 | 6 (4.9) | 0(0.0) | 6 (4.9) | 1 | - |
| Total | 366 | 37 (10.1) | 9 (2.4) | 46 (12.5) | - | - |

OR, odds ratio; 95% CI, confidence interval at 95%.

### Farm practices and risk factors for *Campylobacter* colonization in poultry

In univariable logistic regression, significantly higher *Campylobacter* prevalence was observed in poultry from farms that lacked footbaths or did not change shoes, disposed of poultry waste in the backyard, allowed unauthorized access to the poultry house, failed to separate sick poultry, did not practiced all-in/all-out management, and owned cattle. In multivariable logistic regression, the all-in/all-out management was identified as a protective factor against *Campylobacter* colonization in poultry. Poultry from farms that did not implement all-in/all-out management were 8.4 times (AOR=8.4; 95% CI: 2.2, 32.4) more likely to be colonized by *Campylobacter* compared to poultry from farms that practiced this management, and the difference was statistically significant. Poultry from farms that owned cattle was 9.5 times (AOR=9.5; 95% CI: 1.5, 58.2) more likely to be colonized by *Campylobacter* compared to poultry from farms without cattle (Table 2).

**Table 2.** Univariable (shown as Crude Odds Ratio; COR) and multivariable (shown as Adjusted Odds Ration; AOR) logistic regression analysis of risk factors associated with *Campylobacter* prevalence in cloacal swabs in poultry farms in and around Debre Berhan town (n=122).

| Variables | No. of samples (%) | Positive (%) | COR |  | AOR |  |
| --- | --- | --- | --- | --- | --- | --- |
|  |  |  | 95% CI | P-value | 95% CI | P-value |
| Districts |  |  |  |  |  |  |
| Debre Berhan town | 32 (26.2) | 7 (21.9) | 1.2 (0.4-3.2) | 0.715 |  |  |
| Debre Berhan town surroundings | 90 (73.8) | 17 (18.9) | 1 |  |  |  |
| **Farm location** |  |  |  |  |  |  |
| Urban | 27 (22.1) | 6 (22.2) | 2.1 (0.4-12.1) | 0.388 |  |  |
| Peri-urban | 78 (63.9) | 16 (20.5) | 1.9 (0.4-9.3) | 0.411 |  |  |
| Rural | 17 (13.9) | 2 (11.7) | 1 |  |  |  |
| **Farming system** |  |  |  |  |  |  |
| Small holder | 58 (47.5) | 10 (17.2) | 1 |  |  |  |
| Commercial | 64 (52.5) | 14 (21.8) | 1.3 (0.5-3.3) | 0.521 |  |  |
| **Type of poultry** |  |  |  |  |  |  |
| Broiler | 18 (14.7) | 4 (22.2) | 1.3 (0.3-5.7) | 0.716 |  |  |
| Layer | 70 (57.4) | 13 (18.6) | 1.04 (0.3-3.2) | 0.934 |  |  |
| Mixed | 6 (4.9) | 2 (33.3) | 2.3 (0.3-16.2) | 0.403 |  |  |
| Grower | 28 (22.9) | 5 (17.8) | 1 |  |  |  |
| **Flock size (Average flock size=373)** |  |  |  |  |  |  |
| Small (<20) | 25 (20.5) | 4 (16.0) | 1 |  |  |  |
| Medium (21-99) | 29 (23.8) | 3 (10.3) | 0.6 (0.1-3) | 0.540 |  |  |
| Large (100-999) | 54 (44.2) | 14 (25.9) | 1.8 (0.5-6.3) | 0.332 |  |  |
| Very large (1000-4000) | 14 (11.5) | 3 (21.4) | 1.4 (0.3-7.5) | 0.673 |  |  |
| **Availability of ventilation** |  |  |  |  |  |  |
| Mechanical ventilation | 25 (20.5) | 6 (24.0) | 1 |  |  |  |
| Window | 79 (64.7) | 14 (17.7) | 0.6 (0.23-2) | 0.489 |  |  |
| Under roof | 16 (13.1) | 4 (25.0) | 1.05 (0.2-4.5) | 0.942 |  |  |
| No ventilation | 2 (1.6) | 0 (0.0) | - | - |  |  |
| **Disposal of poultry waste** |  |  |  |  |  |  |
| Backyard | 55 (45.1) | 17 (29.8) | 3.5 (1.3-9.3) | 0.011 | 1.3 (0.32-5.89) | 0.109 |
| Buried, burn or sell | 67 (54.9) | 7 (10.7) | 1 |  |  |  |
| **Use footbaths or change the shoes** |  |  |  |  |  |  |
| Yes | 33 (27.1) | 2 (6.0) | 1 |  |  |  |
| No | 89 (72.9) | 22 (24.7) | 5.1 (1.1-23) | 0.035 | 3.8 (0.62-23.6) | 0.148 |
| **Rodent and insects control** |  |  |  |  |  |  |
| Yes | 51 (41.8) | 9 (17.6) | 1 |  |  |  |
| No | 71 (58.2) | 15 (21.1) | 1.2 (0.5-3.1) | 0.634 |  |  |
| **Drinker and feeder disinfection** |  |  |  |  |  |  |
| Yes | 52 (42.6) | 8 (15.4) | 1 |  |  |  |
| No | 70 (57.4) | 16 (22.8) | 1.6 (0.9-5.9) | 0.307 |  |  |
| **Apply All-In-All-Out management** |  |  |  |  |  |  |
| Yes | 83 (68.1) | 5 (6.0) | 1 |  |  |  |
| No | 39 (31.9) | 19 (48.7) | 14.8 (4.9-44.5) | 0.000 | 8.4 (2.2-32.4) | 0.002 |
| **Restricted access to the flock** |  |  |  |  |  |  |
| Yes | 70 (57.4) | 8 (11.4) | 1 |  |  |  |
| No | 52 (42.6) | 16 (30.7) | 3.4 (1.3-8.8) | 0.010 | 2.5 (0.6-10.7) | 0.210 |
| **Separate sick poultry in the farm** |  |  |  |  |  |  |
| Yes | 66 (54.1) | 8 (12.1) | 1 | 0.026 | 1.4 (0.4-5.5) | 0.627 |
| No | 56 (45.9) | 16 (28.6) | 2.9 (1.1-7.4) |  |  |  |
| **Handling of dead poultry** |  |  |  |  |  |  |
| Throw to garbage<sup>e</sup> | 89 (72.9) | 21 (23.6) | 3 (0.8-11.1) | 0.085 | 1.3(0.2-7.4) | 0.735 |
| Buried or burn | 33 (27.1) | 3 (9.1) | 1 |  |  |  |
| **Type of Confinement** |  |  |  |  |  |  |
| Indoor part time | 48 (39.3) | 16 (33.3) | 1.4 (0.5-3.8) | 0.414 |  |  |
| Indoor exclusively | 70 (57.4) | 8 (29.6) | 1 |  |  |  |
| Outdoor | 4 (3.3) | 0 (0.0) |  |  |  |  |
| **Source of water** |  |  |  |  |  |  |
| Tap water | 74 (60.7) | 19 (25.6) | 2.8 (1.0-8.4) | 0.051 | 3.7 (0.7-18.5) | 0.109 |
| River | 46 (37.7) | 5 (10.8) | 1 |  |  |  |
| Well | 2 (1.6) | 0 (0.0) | - |  |  |  |
| **Presence of other animal species on the farm** |  |  |  |  |  |  |
| No | 41 (33.6) | 2 (4.8) | 1 |  |  |  |
| Cattle | 50 (40.9) | 18 (36.0) | 10.9 (2.3-50.8) | 0.002 | 9.5 (1.5-58.2) | 0.014 |
| Pet (dog and cat) | 8 (6.6) | 1 (12.5) | 2.7 (0.22-35.0) | 0.428 | 9.9 (0.4-221) | 0.146 |
| Small ruminant | 15 (12.3) | 2 (13.3) | 3 (0.38-23.5) | 0.295 | 3.5 (0.2-44.5) | 0.333 |
| Swine | 8 (6.6) | 1 (12.5) | 2.7 (0.22-35) | 0.428 | 4.6 (0.2-139) | 0.372 |
| **Campylobacter positivity in human stool on the farm** |  |  |  |  |  |  |
| Positive | 16 (13.1) | 4 (25.0) | 1.4 (0.41-4.91) | 0.570 |  |  |
| Negative | 106 (86.9) | 20 (18.8) | 1 |  |  |  |
| **Campylobacter contamination status of poultry house floors** |  |  |  |  |  |  |
| Positive | 6 (4.9) | 3 (50.0) | 4.5 (0.85-24) | 0.076 | 1.04 (0.1-11.0) | 0.975 |
| Negative | 116 (95.1) | 21 (18.1) | 1 |  |  |  |

Hosmer-Lemeshow $\chi^2 = 13.36$ , $P=0.100$ ; ROC curve = 0.901 (90%).

### Human consumption habits and hygiene practices

Two-thirds (64.0%) of respondents reported consuming raw meat from ruminant, mainly beef. In a univariable logistic regression, significantly higher prevalence rates were found among individuals with no formal education, farm attendants, those who slaughtered animals at home, individuals who did not use personal protective equipment (PPE) when handling poultry, those who did not wash their hands with soap after contact with live animals, those who disposed of poultry waste using a shovel, and individuals unaware of zoonotic diseases (Table 3). However, in a multivariable logistic regression analysis, it was found that individuals who did not use PPE when handling poultry had 8.3 times (AOR=8.3; 95% CI: 1.9, 36.5) higher odds of contracting *Campylobacter,* and not washing hands after contact with live animals and cleaning barn increased the odds by 5.6 times (AOR=5.6; 95% CI: 1.4, 21.8). Consuming raw dairy products raised the odds by 5.5 times (AOR=5.5; 95% CI:1.0, 29.9), and allowing poultry in human sleeping and food preparation area increased the odds by 6.3 times (AOR=6.3; CI: 1.3, 30.2) (Table 3).

**Table 3.** Univariable (shown as Crude Odds Ratio; COR) and multivariable (shown as Adjusted Odds Ration; AOR) logistic regression analysis of risk factors for *Campylobacter* infection in humans from poultry farms in and around Debre Berhan Town, Ethiopia (n=122).

| Variables | No. of samples (%) | Positive (%) | COR |  | AOR |  |
| --- | --- | --- | --- | --- | --- | --- |
|  |  |  | 95% CI | P-value | 95% CI | P-value |
| Gender |  |  |  |  |  |  |
| Male | 68 (55.7) | 10 (14.7) | 1.4 (0.4-4.1) | 0.469 |  |  |
| Female | 54 (44.3) | 6 (11.1) | 1 |  |  |  |
| Age (*Mean age: 27 years*) |  |  |  |  |  |  |
| ≤20 years | 27 (22.1) | 3 (11.1) | 1 |  |  |  |
| 21-30 years | 55 (45.1) | 7 (12.7) | 1.1 (0.3-4.9) | 0.834 |  |  |
| 31-50 years | 40 (32.7) | 6 (15.0) | 1.4 (0.3-6.2) | 0.648 |  |  |
| Marital status |  |  |  |  |  |  |
| Single | 51 (41.8) | 6(11.7) | 1 |  |  |  |
| Married | 71 (58.2) | 10 (14.1) | 1.2 (0.4-3.6) | 0.708 |  |  |
| Level of education |  |  |  |  |  |  |
| No formal education | 32 (26.2) | 11 (34.4) | 7.8 (1.5-39.2) | 0.012 |  |  |
| Primary school | 32 (26.2) | 2 (6.2) | 1 |  |  |  |
| High school | 38 (31.2) | 2 (5.2) | 0.8 (0.1-6.2) | 0.860 |  |  |
| Higher education | 20 (16.4) | 1 (5.0) | 0.8 (0.1-9.3) | 0.851 |  |  |
| **Occupation** |  |  |  |  |  |  |
| Farm attendant | 75 (61.5) | 15 (20.0) | 10.7 (1.4-84.5) | 0.024 |  |  |
| Farm manager | 44 (36) | 1 (2.3) | 1 |  |  |  |
| Other<sup>a</sup> | 3 (3.0) | 0 (0.0) |  |  |  |  |
| **Eat undercooked meat** |  |  |  |  |  |  |
| Yes | 78 (63.9) | 13 (16.7) | 2.7 (0.7-10.1) | 0.134 |  |  |
| No | 44 (36.1) | 3 (6.8) | 1 |  |  |  |
| **Drink raw milk** |  |  |  |  |  |  |
| Yes | 71 (58.1) | 14 (20) | 6 (1.3-27.8) | 0.022 | 5.5 (1.0-29.9) | 0.046 |
| No | 51 (41.8) | 2 (3.9) | 1 |  |  |  |
| **Access poultry to human sleeping and food preparation areas** |  |  |  |  |  |  |
| Yes | 77 (63.1) | 13 (16.9) | 2.8 (0.7-10.6) | 0.119 | 6.3 (1.3-30.2) | 0.020 |
| No | 45 (36.9) | 3 (6.7) | 1 |  |  |  |
| **Slaughter domestic animals at their home** |  |  |  |  |  |  |
| Yes | 69 (56.5) | 13 (18.8) | 3.8 (1.0-14.3) | 0.043 |  |  |
| No | 53 (43.4) | 3 (6.9) | 1 |  |  |  |
| **Use of PPE<sup>b</sup> during contact with poultry** |  |  |  |  |  |  |
| Yes | 71 (58.2) | 3 (4.2) | 1 |  |  |  |
| No | 51 (41.8) | 13 (25.5) | 7.7 (2.1-28.9) | 0.002 | 8.3 (1.9-36.5) | 0.005 |
| **Wash hands with soap after handling live poultry and cleaning barn** |  |  |  |  |  |  |
| Yes | 80 (65.6) | 4 (5.0) | 1 |  |  |  |
| No | 42 (34.4) | 12 (28.6) | 7.6 (2.3-25.4) | 0.001 | 5.6 (1.4-21.8) | 0.013 |
| **How do you dispose the poultry wastes<sup>c</sup>?** |  |  |  |  |  |  |
| Using glove | 35 (28.7) | 1 (2.8) | 1 |  |  |  |
| By bare hand | 65 (53.3) | 9 (13.8) | 5.4 (0.7-45.0) | 0.115 |  |  |
| Using shovel | 22 (18.0) | 6 (27.3) | 12.7 (1.4-44.9) | 0.023 |  |  |
| **Zoonosis Awareness** |  |  |  |  |  |  |
| Yes | 47 (38.5) | 2 (4.2) | 1 |  |  |  |
| No | 75 (61.5) | 14 (18.7) | 5.16 (1.1-23.8) | 0.036 |  |  |
| **Poultry colonized with *Campylobacter* on the farm** |  |  |  |  |  |  |
| Positive | 24 (29.6) | 4 (16.7) | 1.4 (0.4-4.9) | 0.567 |  |  |
| Negative | 98 (80.3) | 12 (12.2) | 1 |  |  |  |
| ***Campylobacter* contamination status of poultry house floors** |  |  |  |  |  |  |
| Positive | 6 (4.9) | 1 (16.7) | 1.3 (0.1-12.3) | 0.792 |  |  |
| Negative | 116 (95.1) | 15 (12.9) | 1 |  |  |  |

<sup>a</sup>Teacher or animal health professional or bankers; <sup>b</sup>PPE, personal protective equipment; wastes<sup>c</sup>,
manure, aborted fetus; Hosmer-Lemeshow chi ( $\chi^2$ ) = 1.01, P = 0.994; ROC curve = 0.872 (87%).

### Awareness of poultry farm workers

The majority of the respondents (61.5%) were unaware of zoonotic diseases. The zoonotic diseases mentioned by the 47 award respondents included tuberculosis (n=13), cholera (n=10), salmonellosis (n=6), anthrax (n=2), and both tuberculosis and cholera (n=5). Eleven mentioned ‘bacteria’ as a zoonotic disease. None of the interviewees were aware of *Campylobacter* or recognized it as a zoonotic disease. Only 5 (4.1%) of respondents were familiar with the One Health concept. Regarding attitudes toward zoonotic disease transmission, 57 (46.7%) believed that diarrhea-causing agents could be transmitted from animals to humans. The routes of transmission identified by these 57 respondents were: consumption of contaminated raw meat (n=13), contaminated milk and meat (n=12), contaminated meat and water (n=8), contaminated raw milk (n=7), contact with infected animals (n=9), exposure to contaminated water (n=4), and unknown (n=4).

### Antimicrobial resistance profiles

All 46 *Campylobacter* isolates exhibited resistance to at least one antimicrobial. Overall, tetracycline showed the highest resistance (89.1%), followed by ciprofloxacin (69.5%) and erythromycin (63.0%). Among the 37 *C. jejuni* isolates, 92.0% were resistant to tetracycline, 70.2% to ciprofloxacin, and 64.8% to erythromycin. For the 9 *C. coli* isolates, resistance rates were 77.8% to tetracycline, 66.7% to ciprofloxacin, and 55.5% to erythromycin. By considering only no inhibition of growth (0 mm), resistance was also observed for other antimicrobials: 28.2% of isolates were resistant to norfloxacin (32.4% *C. jejuni* and 11.1% *C. coli*), 30.4% to streptomycin (29.7% *C. jejuni* and 33.3% *C. coli*), 13.5% to chloramphenicol (*C. jejuni*), 21.7% to gentamicin (24.3% *C. jejuni* and 11.1% *C. coli*), 19.5% to nalidixic acid (18.9% *C. jejuni* and 22.2% *C. coli*), 73.9% to oxytetracycline (78.3% *C. jejuni* and 55.5% *C. coli*) (Table 4).

**Table 4.** Distribution of antimicrobial resistance among 37 *C. jejuni* and 9 *C. coli* isolated from human stool, cloacal swabs, and poultry house floors in and around Debre Berhan, Ethiopia. Grey rows indicate MDR isolates, i.e. resistant to at least three different antibiotic classes.

| *C. jejuni* resistance | Human<br>(n=14) | Cloacal<br>(n=17) | Sock samples<br>(n=6) | Total |
| --- | --- | --- | --- | --- |
| Cip+Ery+Tet | 3 |  |  | 3 |
| Cip+Ery+Tet+Oxy* | 1 | 2 | 1 | 4 |
| Cip+Ery+Tet+Oxy*+Nor* | 1 | 2 | 1 | 4 |
| Cip+Ery+Tet+Oxy*+Nor*+Str* | 1 | 1 |  | 2 |
| Cip+Ery+Tet+Oxy*+Nor*+Str*+Nal* | 1 | 1 | 1 | 3 |
| Cip+Ery+Tet+Oxy*+Nor*+Gen*+Nal* |  | 1 | 1 | 2 |
| Cip+Ery+Tet+Oxy*+Gen* |  | 3 |  | 3 |
| Cip+Tet+Oxy*+Str*+Gen* | 1 |  |  | 1 |
| Cip+Ery+Tet+Oxy*+Str*+Gen*+Chl* | 1 |  |  | 1 |
| Cip+Ery+Tet+Oxy*+Nor*+Gen*+Str*+Chl* |  | 1 |  | 1 |
| Ery+Tet+Oxy*+Str* |  | 2 |  | 2 |
| Ery+Tet+Oxy*+Chl* |  | 1 | 1 | 2 |
| Ery+Tet+Oxy* |  |  | 1 | 1 |
| Ery+Tet | 2 |  |  | 2 |
| Cip+Tet+Oxy* | 1 |  |  | 1 |
| Tet+Oxy* |  | 2 |  | 2 |
| Ery | 2 |  |  | 2 |
| Str* |  | 1 |  | 1 |
| ***C. coli* resistance** | **Human**<br>**(n=2)** | **Cloacal**<br>**(n=7)** | **Sock samples**<br>**(n=0)** |  |
| Cip+Ery+Tet+Oxy* | 1 | 1 |  | 2 |
| Cip+Ery+Tet+Oxy*+Str*+Gen*+Chl* |  | 1 |  | 1 |
| Cip+Ery+Tet+Oxy*+Str*+Nor* |  | 1 |  | 1 |
| Cip+Nal* |  | 1 |  | 1 |
| Cip+ Nal*+Tet+Oxy* |  | 1 |  | 1 |
| Str* |  | 1 |  | 1 |
| Ery | 1 |  |  | 1 |
| Gen* |  | 1 |  | 1 |

\*No validated cut-off value is available, resistance defined as absence of growth inhibition (0 mm).
Abbreviations: Cip, Ciprofloxacin; Ery, Erythromycin; Tet, Tetracycline; Oxy, Oxytetracycline; Nor, Norfloxacin; Str, Streptomycin; Nal, Nalidixic acid; Gen, Gentamicin and Chl, Chloramphenicol.

Overall, 69.5% (32/46) of the *Campylobacter* isolates were MDR, with 75.7% (28/37) of *C. jejuni* and 44.4% (4/9) of *C. coli* strains. Among the 28 MDR *C. jejuni* isolates, 82.3% (14/17) were from poultry, 64.3% (9/14) from humans, and 83.3% (5/6) from poultry house floors. The four MDR *C. coli* isolates originated from three poultry samples and one human sample. The most common MDR pattern involved resistance to quinolones, macrolides, and tetracyclines (Table 4). All co-occurring *C. jejuni* strains on the same farms were MDR, with similar resistance patterns observed across all sample types on five farms, except for isolates from one farm.

## Discussion

The occurrence of *Campylobacter* in this study aligns with previous research from Ethiopia [12, 27, 28], Kenya [29], Sub-Saharan Africa [30], and Cambodia [31]. However, our findings show a lower prevalence than earlier studies in Ethiopia [32–34] and Tanzania [35], likely due to differences in geography, detection methods, sampling protocols, animal management, and sanitation practices.

In this study, *Campylobacter* was detected in 4.9% of poultry floor samples, highlighting fecal contamination as a key factor in the spread of *Campylobacter*. A notably higher prevalence of *Campylobacter* (18.4%) has been reported in a previous study conducted in Ethiopia [36]. This discrepancy may reflect a genuine difference in prevalence rates or could be attributed to variations in detection methodologies. Specifically, the aforementioned study utilized direct polymerase chain reaction (PCR) techniques on environmental samples, whereas our investigation employed culture methods with enrichment using buffer peptone water without the addition of antibiotic supplement to inhibit microorganisms such as coliform bacteria, yeast and molds, which were subsequently confirmed by PCR. This methodological divergence may account for the lower prevalence observed in our study.

*Campylobacter jejuni* was the dominant species isolated in this study, similar to previous studies [30–35], highlighting its critical role in human campylobacteriosis [10]. The co-occurrence of *C. jejuni* across various sample types within the same farm, including poultry, humans, and the environment, suggests complex transmission dynamics at the farm level. This assertion is further supported by the observation of similar AMR patterns across these isolates, all of which were MDR, indicating a potential transmission cycle within the farm environment.

Our study found that the all-in/all-out management system significantly reduces *Campylobacter* colonization in poultry. The absence of this system in some farms highlights a critical biosecurity gap that contributes to the spread of *Campylobacter*. The all-in/all-out approach minimizes cross-contamination between flocks and facilitates thorough cleaning between batches, thus reducing bacterial loads. Inadequate cleaning, partial depopulations, and poor disinfection practices between batches were identified as a risk factors for *Campylobacter* colonization [37]. Additionally, farms that housed both poultry and cattle were found to have higher odds of having *Campylobacter*-positive poultry, suggesting horizontal transmission between cattle and poultry. Previous studies have consistently identified other animals, particularly cattle, on or near farms as significant risk factors [6,38], likely playing a role in maintaining *Campylobacter* spp. between flocks [37]. Therefore, other domestic animal species could play a role in *Campylobacter* transmission to humans, especially in countries such as Ethiopia, where mixed livestock farming is a common practice.

This study also identified that raw milk consumption was a risk factor for human *Campylobacter* infection, consistent with findings from previous studies [39, 40]. In addition, allowing poultry to roam in human living and food preparation areas significantly increased the likelihood of infection, similar to findings from Cambodia [31]. Poor hygiene practices, such as failure to wash hands after handling poultry or cleaning poultry barns, and not using PPE, were also associated with higher infection rates. These results reinforce the importance of hygiene as a major risk factor for *Campylobacter* transmission, as highlighted in previous studies [27, 41].

A significant knowledge gap was observed among farm workers regarding zoonotic diseases and the One Health concept, with all workers unaware that *Campylobacter* is zoonotic. This aligns with findings from Abunna et al. [42], which also identified limited awareness of zoonotic risks as a barrier to improving public health outcomes. Our univariable analysis showed that *Campylobacter* prevalence was higher in individuals unaware of its zoonotic nature, likely due to their lack of awareness. These findings underscore the need for targeted educational interventions to raise awareness about zoonotic risks and promote One Health strategies to reduce risks to both animal and human health [43].

Antimicrobial resistance was a major concern in this study. All *Campylobacter* isolates showed resistance to at least one antimicrobial agent, with high resistance levels to tetracycline, erythromycin, and ciprofloxacin. These findings are consistent with previous studies from Ethiopia [12, 27], other Sub-Saharan African countries [30] and globally [8]. Such high resistance is likely driven by the widespread use of antibiotics as prophylactics, with nearly half of the poultry farms in the study area relying on them. Tetracyclines, especially oxytetracycline, have long been used in Ethiopian animal production. Improper antimicrobial use in poultry farming, coupled with insufficient regulatory oversight, has contributed to the emergence of resistant bacterial strains [12, 44, 45]. The long-term, uncontrolled use of fluoroquinolones and tetracycline, coupled with poor regulatory oversight, has facilitated the spread of resistance [11].

The high MDR rate observed in this study is a serious health issue, as most of the isolates were resistant to commonly used antibiotics. The true MDR rate could be much higher, as in this study only no inhibition of growth (0 mm) were considered resistant for certain antibiotics. Similar findings have been reported from Ethiopia [27, 46, 47], Kenya [29], and Bangladesh [48]. The dominant MDR pattern includes resistance to quinolones, macrolides, and tetracyclines, reflecting global trends of high resistance, particularly to tetracycline and ciprofloxacin [49]. The co-occurrence of MDR isolates on farms underscores the emergence and transmission of antibiotic-resistant *Campylobacter*. Bidirectional transmission between humans and animals, coupled with widespread antimicrobial treatments, leads to continuous antibiotic pressure, facilitating the increase and spread of MDR *Campylobacter* [47, 50].

## Conclusion

In conclusion, this study highlights the widespread occurrence of *Campylobacter* in poultry and humans, along with antibiotic resistance. Poor biosecurity and management practices contributed to poultry colonization, while human infections were mostly linked to consumption of raw milk and inadequate hygiene practices on poultry farms. A knowledge gap among farm workers underscores the need for targeted education. These findings emphasize the value of a One Health approach. Future interventions in Ethiopia should focus on strengthening biosecurity, improving hygiene, and implementing antimicrobial stewardship programs to reduce risks to both animal and human health.

## Data Availability

All data will be available at Swedish University of Agriculture Sciences (SLU) repository.

## Acknowledgments

We acknowledge Dr. Wondwesen Azemeraw for assisting with data collection and Dr. Mequanint Addisu for his support with laboratory work. We also appreciate the poultry farm owners, district authorities, veterinarians, and field workers who facilitated and supported the fieldwork.

## References

1. Li M, Havelaar AH, Hoffmann S, Hald T, Kirk MD, Torgerson PR, et al. Global disease burden of pathogens in animal source foods, 2010. PLoS ONE. 2019;14(6):e0216545. doi: 10.1371/journal.pone.0216545.

2. Costa D, Iraola G. Pathogenomics of emerging *Campylobacter* species. Clin Microbiol Rev. 2019 Jul 3;32(4):e00072–18. doi: 10.1128/CMR.00072-18. PMID: 31270126; PMCID: PMC6750134.

3. Sahin O, Yaeger M, Wu Z, Zhang Q. *Campylobacter*-Associated Diseases in Animals. Annu Rev Anim Biosci. 2017 Feb 8;5:21–42. doi: 10.1146/annurev-animal-022516-022826. Epub 2016 Nov 9. PMID: 27860495.

4. Kaakoush NO, Castaño-Rodríguez N, Mitchell HM, Man SM. Global epidemiology of *Campylobacter* infection. Clin Microbiol Rev. 2015 Jul;28(3):687–720. doi: 10.1128/CMR.00006-15. PMID: 26062576; PMCID: PMC4462680.

5. Conrad CC, Stanford K, Narvaez-Bravo C, Callaway T, McAllister T. Farm fairs and petting zoos: A review of animal contact as a source of zoonotic enteric disease. Foodborne Pathog Dis. 2017 Feb;14(2):59–73. doi: 10.1089/fpd.2016.2185. Epub 2016 Dec 19. PMID: 27992253.

6. Hansson I, Olsson Engvall E, Vågsholm I, Nyman A. Risk factors associated with the presence of *Campylobacter*-positive broiler flocks in Sweden. Prev Vet Med. 2010;96(1-2):114–121. doi: 10.1016/j.prevetmed.2010.05.007.

7. Sommer HM, Heuer OE, Sørensen AI, Madsen M. Analysis of factors important for the occurrence of *Campylobacter* in Danish broiler flocks. Prev Vet Med. 2013 Aug 1;111(1-2):100–11. doi: 10.1016/j.prevetmed.2013.04.004. Epub 2013 May 21. PMID: 23706344.

8. Van Boeckel TP, Pires J, Silvester R, Zhao C, Song J, Criscuolo NG, et al. Global trends in antimicrobial resistance in animals in low- and middle-income countries. Science. 2019 Sep 20;365(6459):eaaw1944. doi: 10.1126/science.aaw1944. PMID: 31604207.

9. Tacconelli E, Carrara E, Savoldi A, Harbarth S, Mendelson M, Monnet DL, et al. Discovery, research, and development of new antibiotics: the WHO priority list of antibiotic-resistant bacteria and tuberculosis. Lancet Infect Dis. 2018 Mar;18(3):318–327. doi: 10.1016/S1473-3099(17)30753-3. Epub 2017 Dec 21. PMID: 29276051.

10. Zhang Q, Beyi AF, Yin Y. Zoonotic and antibiotic-resistant *Campylobacter*: a view through the One Health lens. One Health Adv. 2023;1:4. doi: 10.1186/s44280-023-00003-1.

11. Elhadidy M, Miller WG, Arguello H, Álvarez-Ordóñez A, Duarte A, Dierick K, et al. Genetic basis and clonal population structure of antibiotic resistance in *Campylobacter jejuni* isolated from broiler carcasses in Belgium. Front Microbiol. 2018 May 16;9:1014. doi: 10.3389/fmicb.2018.01014.

12. Zenebe T, Zegeye N, Eguale T. Prevalence of *Campylobacter* species in human, animal, and food of animal origin and their antimicrobial susceptibility in Ethiopia: a systematic review and meta-analysis. Ann Clin Microbiol Antimicrob. 2020 Dec 10;19(1):61. doi: 10.1186/s12941-020-00405-8. PMID: 33302968; PMCID: PMC7731538.

13. Central Statistical Agency (CSA). Federal Democratic Republic of Ethiopia, Central Statistical Agency (CSA) Agricultural sample survey, Volume II, report on livestock and livestock characteristics (private peasant holdings). Statistical bulletin 589. Addis Ababa, Ethiopia; March 2021.

14. Food and Agriculture Organization (FAO). Poultry Sector Ethiopia: FAO Animal Production and Health Livestock Country Reviews No. 11. Rome: Food and Agriculture Organization; 2019. Accessed 16 September 2024. Available from: https://books.google.se/books?id=FC6rDwAAQBAJ.

15. Climate: Debre Berhan. Retrieved 10 September 2024 from https://www.meteoblue.com/en/weather/historyclimate/climatemodelled/debre-birhan_ethiopia_339734.

16. Thrusfield M. Veterinary Epidemiology. 3rd ed. Wiley; 2007. ISBN: 1405156279, 9781405156271. 624 pages. ISBN: 1405156279, 9781405156271.

17. Hansson I, Vågsholm I, Svensson L, Olsson Engvall E. Correlations between *Campylobacter spp*. prevalence in the environment and broiler flocks. J Appl Microbiol. 2007 Sep;103(3):640–9. doi: 10.1111/j.1365-2672.2007.03291.x. PMID: 17714397.

18. ISO. International Organization for Standardization. ISO 10272 Microbiology of Food and Animal Feeding Stuff—Horizontal Method for Detection and Enumeration of Campylobacter spp.—Part 1: Enrichment Method. ISO; Geneva, Switzerland: 2017.

19. Oyarzabal OA, Backert S, Nagaraj M, Miller SR, Hussain KS, Oyarzabal AE. Efficacy of supplemented buffered peptone water for the isolation of *Campylobacter jejuni* and *C. coli* from broiler retail products. J Microbiol Methods. 2007;69(1):129–36. doi: 10.1016/j.mimet.2006.12.011.

20. Barakat AMA, El-Razik KAA, Elfadaly HA, Rabie NS, Sadek SAS, Almuzaini AM. Prevalence, molecular detection, and virulence gene profiles of *Campylobacter* species in humans and foods of animal origin. Veterinary World. 2020; 13(7): 1430–1438. 10.14202/vetworld.2020.1430-1438.

21. Linton D, Lawson AJ, Owen RJ, Stanley J. PCR detection, identification to species level, and fingerprinting of *Campylobacter jejuni* and *Campylobacter coli* direct from diarrheic samples. J Clin Microbiol. 1997 Oct;35(10):2568–72. doi: 10.1128/jcm.35.10.2568-2572.1997. PMID: 9316909; PMCID: PMC230012.

22. Denis M, Soumet C, Rivoal K, Ermel G, Blivet D, Salvat G, Colin P. Development of a m-PCR assay for simultaneous identification of *Campylobacter jejuni* and *Campylobacter coli*. Lett Appl Microbiol. 1999 Dec;29(6):406–10. doi: 10.1046/j.1472-765x.1999.00658.x. PMID: 10664985.

23. Wang G, Clark CG, Taylor TM, Pucknell C, Barton C, Price L, Woodward DL, Rodgers FG. Colony multiplex PCR assay for identification and differentiation of *Campylobacter jejuni, C. coli, C. lari*, C. upsaliensis, and C. fetus subsp. fetus. J Clin Microbiol. 2002 Dec;40(12):4744–7. doi: 10.1128/JCM.40.12.4744-4747.2002. PMID: 12454184; PMCID: PMC154608.

24. Yamazaki-Matsune W, Taguchi M, Seto K, Kawahara R, Kawatsu K, Kumeda Y, et al. Development of a multiplex PCR assay for identification of *Campylobacter coli, Campylobacter fetus, Campylobacter hyointestinalis* subsp. *hyointestinalis, Campylobacter jejuni, Campylobacter lari* and *Campylobacter upsaliensis*. J Med Microbiol. 2007 Nov;56(Pt 11):1467–1473. doi: 10.1099/jmm.0.47363-0. PMID: 17965346.

25. Banowary B, Dang VT, Sarker S, Connolly JH, Chenu J, Groves P, et al. Differentiation of *Campylobacter jejuni* and *Campylobacter coli* Using Multiplex-PCR and High Resolution Melt Curve Analysis. PLoS One. 2015 Sep 22;10(9):e0138808. doi: 10.1371/journal.pone.0138808. PMID: 26394042; PMCID: PMC4578860.

26. The European Committee on Antimicrobial Susceptibility Testing. Breakpoint tables for interpretation of MICs and diameters zones. Version 15, 2025. Available at http://www.eucast.org/.

27. Chala G, Eguale T, Abunna F, Asrat D, Stringer A. Identification and Characterization of *Campylobacter* Species in Livestock, Humans, and Water in Livestock Owning Households of Peri-urban Addis Ababa, Ethiopia: A One Health Approach. Front Public Health. 2021;9:750551. doi: 10.3389/fpubh.2021.750551.

28. Belina D, Gobena T, Kebede A, Chimdessa M, Mummed B, Thystrup CAN, et al. Occurrence and diversity of *Campylobacter* species in diarrheic children and their exposure environments in Ethiopia. PLOS Glob Public Health. 2024;4(10):e0003885. doi: 10.1371/journal.pgph.0003885.

29. Abubakar MK, Muigai AWT, Ndung’u P, Kariuki S. Investigating carriage, contamination, antimicrobial resistance and assessment of colonization risk factors of *Campylobacter* spp. in broilers from selected farms in Thika, Kenya. Microbiol Res J Int. 2019;27:1–16. 10.9734/mrji/2019/v27i630119.

30. Gahamanyi N, Mboera LE, Matee MI, Mutangana D, Komba EV, et al. Prevalence, Risk Factors, and Antimicrobial Resistance Profiles of Thermophilic *Campylobacter* Species in Humans and Animals in Sub-Saharan Africa: A Systematic Review. Int J Microbiol. 2020;2020: Article ID 2092478.

31. Osbjer K, Boqvist S, Sokerya S, Chheng K, San S, Davun H, et al. Risk factors associated with *Campylobacter* detected by PCR in humans and animals in rural Cambodia. Epidemiol Infect. 2016 Oct;144(14):2979–2988. doi: 10.1017/S095026881600114X. Epub 2016 Jun 23. PMID: 27334412; PMCID: PMC5080667.

32. Kassa T, Gebre-Selassie S, Asrat D. Antimicrobial susceptibility patterns of thermotolerant *Campylobacter* strains isolated from food animals in Ethiopia. Vet Microbiol. 2007 Jan 17;119(1):82–7. doi: 10.1016/j.vetmic.2006.08.011. Epub 2006 Aug 14. PMID: 17000061.

33. Budge S, Barnett M, Hutchings P, Parker A, Tyrrel S, Hassard F, et al. Risk factors and transmission pathways associated with infant *Campylobacter* spp. prevalence and malnutrition: A formative study in rural Ethiopia. PLoS ONE. 2020;15(5):e0232541. 10.1371/journal.pone.0232541.

34. Mulu W, Joossens M, Kibret M, Van den Abeele A-M, Houf K. *Campylobacter* occurrence and antimicrobial resistance profile in under five-year-old diarrheal children, backyard farm animals, and companion pets. PLoS Negl Trop Dis. 2024;18(6):e0012241. 10.1371/journal.pntd.0012241.

35. Chuma IS, Nonga HE, Mdegela RH, Kazwala RR. Epidemiology and RAPD-PCR typing of thermophilic campylobacters from children under five years and chickens in Morogoro Municipality, Tanzania. BMC Infect Dis. 2016 Nov 21;16(1):692. doi: 10.1186/s12879-016-2031-z. PMID: 27871251; PMCID: PMC5117500.

36. Brena MC, Mekonnen Y, Bettridge JM, Williams NJ, Wigley P, Sisay TT, et al. Changing risk of environmental *Campylobacter* exposure with emerging poultry production systems in Ethiopia. Epidemiol Infect. 2016;144(3):567–575. doi: 10.1017/S0950268815001429.

37. EFSA BIOHAZ Panel (EFSA Panel on Biological Hazards), Koutsoumanis K, Allende A, Alvarez-Ordonez A, Bolton D, Bover-Cid S, et al. Update and review of control options for *Campylobacter* in broilers at primary production. EFSA Journal. 2020;18(4):6090, 89 pp. doi: 10.2903/j.efsa.2020.609.

38. Ellis-Iversen J, Jorgensen F, Bull S, Powell L, Cook AJ, Humphrey TJ. Risk factors for *Campylobacter* colonization during rearing of broiler flocks in Great Britain. Prev Vet Med. 2009 Jun 1;89(3-4):178–84. doi: 10.1016/j.prevetmed.2009.02.004. Epub 2009 Mar 28. PMID: 19329201.

39. Diriba K, Awulachew E, Anja A. Prevalence and associated factor of *Campylobacter* species among less than 5-year-old children in Ethiopia: a systematic review and meta-analysis. Eur J Med Res. 2021 Jan 3;26(1):2. doi: 10.1186/s40001-020-00474-7. PMID: 33390175; PMCID: PMC7780653.

40. Lahti E, Rehn M, Ockborn G, Hansson I, Ågren J, Engvall EO, Jernberg C. Outbreak of Campylobacteriosis Following a Dairy Farm Visit: Confirmation by Genotyping. Foodborne Pathog Dis. 2017 Jun;14(6):326–332. doi: 10.1089/fpd.2016.2257. Epub 2017 Mar 28. PMID: 28350214.

41. Conan A, O’Reilly CE, Ogola E, Ochieng JB, Blackstock AJ, Omore R, Ochieng L, Moke F, Parsons MB, Xiao L, et al. Animal-related factors associated with moderate-to-severe diarrhea in children younger than five years in western Kenya: A matched case-control study. PLoS Negl Trop Dis. 2017;11:e0005795. doi: 10.1371/journal.pntd.0005795.

42. Abunna F, Gebresenbet G, Megersa B. Assessment of knowledge, attitude and practices (KAP) of farmers about transmission of zoonotic diseases in Ada’a district, Oromia, Ethiopia. Heliyon. 2024 Feb 8;10(4):e25713. doi: 10.1016/j.heliyon.2024.e25713. PMID: 38384538; PMCID: PMC10878875.

43. Boqvist S, Söderqvist K, Vågsholm I. Food safety challenges and One Health within Europe. Acta Vet Scand. 2018 Jan 3;60(1):1. doi: 10.1186/s13028-017-0355-3. PMID: 29298694; PMCID: PMC5751857.

44. Abdi RD, Mengstie F, Beyi AF, Beyene T, Waktole H, Mammo B, et al. Determination of the sources and antimicrobial resistance patterns of *Salmonella* isolated from the poultry industry in Southern Ethiopia. BMC Infect Dis. 2017 May 18;17(1):352. doi: 10.1186/s12879-017-2437-2. PMID: 28521744; PMCID: PMC5437651.

45. Gemeda BA, Amenu K, Magnusson U, Dohoo I, Hallenberg GS, Alemayehu G, et al. Antimicrobial use in extensive smallholder livestock farming systems in Ethiopia: Knowledge, attitudes, and practices of livestock keepers. Front Vet Sci. 2020 Feb 26;7:55. doi: 10.3389/fvets.2020.00055. PMID: 32175334; PMCID: PMC7055293.

46. Abamecha A, Assebe G, Tafa B, Wondafrash B. Prevalence of thermophilic *Campylobacter* and their antimicrobial resistance profile in food animals in Lare District, Nuer Zone, Gambella, Ethiopia. J Drug Res Dev. 2015;1(2):108. doi: 10.16966/jdrd.108

47. Debelo M, Mohammed N, Tiruneh A, Tolosa T. Isolation, identification and antibiotic resistance profile of thermophilic *Campylobacter* species from bovine, knives, and personnel at Jimma Town Abattoir, Ethiopia. PLoS ONE. 2022;17(10):e0276625. 10.1371/journal.pone.0276625.

48. Neogi SB, Islam MM, Islam SKS, Akhter AHMT, Sikder MMH, Yamasaki S, Kabir SML. Risk of multi-drug-resistant *Campylobacter* spp. and residual antimicrobials at poultry farms and live bird markets in Bangladesh. BMC Infect Dis. 2020 Apr 15;20(1):278. doi: 10.1186/s12879-020-05006-6. PMID: 32293315; PMCID: PMC7158023.

49. Signorini M, Rossler E, Díaz DC, Olivero C, Romero-Scharpen A, Soto L, et al. Antimicrobial resistance of thermotolerant *Campylobacter* species isolated from humans, food-producing animals, and products of animal origin: A worldwide meta-analysis. Microb Drug Resist. 2018 Oct 1. doi: 10.1089/mdr.2017.0310. Corpus ID: 20883102.

50. Qin X, Wang X, Shen Z. The rise of antibiotic resistance in *Campylobacter*. Curr Opin Gastroenterol. 2023 Jan 1;39(1):9–15. doi: 10.1097/MOG.0000000000000901. Epub 2022 Nov 11. PMID: 36504031.

